# Evaluating risk detection methods to uncover ontogenic-mediated adverse drug effect mechanisms in children

**DOI:** 10.1101/2021.03.10.21253302

**Authors:** Nicholas P. Giangreco, Nicholas P. Tatonetti

**Affiliations:** Departments of Systems Biology and Biomedical Informatics, Columbia University, 622 W. 168^th^ Street, New York, NY, 10032

**Keywords:** Pharmacovigilance, Pediatrics, Child Development, Modeling, Dynamics

## Abstract

**Background:** Identifying adverse drugs effects (ADEs) in children is essential for preventing disability and death from marketed drugs. At the same time, however, detection is challenging due to dynamic biological processes during growth and maturation, called ontogeny, that alter pharmacokinetics and pharmacodynamics. As a result, current data mining methodologies have been limited to event surveillance and have not focused on investigating adverse event mechanisms. There is an opportunity to design data mining methodologies to identify and evaluate drug event patterns within observational databases for ontogenic-mediated adverse event mechanisms. The first step of which is to establish statistical models that can identify temporal trends of adverse effects across childhood.

**Results:** Using simulation, we evaluated a population stratification method (the proportional reporting ratio or PRR) and a population modeling method (the generalized additive model or GAM) to identify and quantify ADE risk at varying reporting rates and dynamics. We found that GAMs showed improved performance over the PRR in detecting dynamic drug event reporting across child developmental stages. Moreover, GAMs exhibited normally distributed and robust ADE risk estimation at all development stages by sharing information across child development stages.

**Conclusions:** Our study underscores the opportunity for using population modeling techniques, which leverages drug event reporting across development stages, to identify adverse drug effect risk resulting from ontogenic mechanisms.

## Background

Adverse drug events (ADEs) in children are common and can result in injury and death^1,2^. Clinical trials rarely include children^3^ and pediatric-specific trials are limited in identifying possible ADEs in the population^4^. Pediatric drug safety studies can evaluate large numbers of ADEs from the population^5^ but current methodologies are limited in their ability to identify the mechanisms that drive pediatric ADEs^6^. Children undergo evolutionarily conserved and physiologically dynamic biological processes, collectively called ontogeny, as they grow and develop from birth through adolescence^7,8^. The mechanisms may include varying protein activity^9,10^ as well as include functional and structural changes that occur during maturation^11,12^. These ontogenic changes can alter pharmacodynamics and pharmacokinetics resulting in adverse effects, as is the case for doxorubicin-induced cardiotoxicity^13^ and valproate-induced hepatotoxicity^14^. With a few notable exceptions, however, many pediatric adverse events are idiopathic with no known, clear connection to developmental biology^15,16^. Additionally, adverse event mechanisms established in adults may not translate to the pediatric population^17^. There is an opportunity to combine known ontogenic biology with real-world pediatric drug effect data to identify ontogenic-mediated adverse events.

To date, elucidation of ontogenic mechanisms has relied on hypothesis-driven approaches. For example, juvenile mouse models have been used to identify genetic vulnerabilities of hematopoiesis^18^ and investigate effects by a glutamatergic agonist on the neural developmental sequence^19^ during early life. More recently, pharmacometric tools have been used to extrapolate drug effects from adults to children, such as projecting acetaminophen exposure across pediatric age groups^20^, and investigate drug action in children, such as predicting clearance of zidovudine during infancy^21^. However, juvenile animal studies are low-throughput and require complex study designs^22^, and there is limited experimental data to parameterize manually designed pharmacometric models^23,24^. While lacking specificity, top-down studies are complementary in that they evaluate thousands of hypotheses simultaneously and can identify idiosyncratic effects that would otherwise go unnoticed^25,26^. Moreover, analyses of large population datasets start from clinically significant events which can take decades to identify^27,28^. Top-down studies can close the pediatric evidence gap^23^ by sifting through large databases to identify clinically significant although perhaps less studied and rare adverse drug events during the period of child growth and development.

While pediatric pharmacovigilance has been able to identify adverse drug events, it is limited in identifying growth and development processes that underlie those observations^10,29^. A common approach when identifying ADEs is to stratify the pediatric population into age groups which directly reduces the amount of data available to identify ADEs during childhood. The Proportional Reporting Ratio, which was designed to be sensitive even when data is scarce^30^, is an established detection method and has been shown to unmask ADE signal within child development stages compared to detection within the larger pediatric population^31^. However, reduced data within these strata was shown to significantly affect PRR detection performance across pediatric age groups^31^. To investigate pediatric ADEs, the continuous, time-dependent biological processes during growth and development suggest using all information across child development stages.

Generalized additive models (GAMs) are supervised machine learning approaches that can quantify non-linear effects reflective of natural phenomena^32^. GAMs may be able to quantify signal reflecting dynamic, continuous processes such as ontogeny. These models are extensively used for spatial and temporal analysis in ecological studies^33^, such as explaining cardiovascular mortality risk from heat waves over time^34^ and rat infestation from environmental factors within geographic areas^35^. Similar to evaluating ecological responses using shared information across time or space, we can evaluate adverse events from temporally-connected ontogenic processes using shared information across child development stages.

We performed the first study to directly evaluate dynamic drug event reporting during childhood. We performed a data simulation and augmentation study that 1) simulated drug event reporting temporal trends of different effect sizes and shapes, 2) augmented existing pediatric drug event data by inserting the simulated reporting rates within observational data, and 3) evaluated population stratification (PRR) and modeling (GAM) methods to detect these injected ADE reporting dynamics. We found the detection scores generated by the GAM showed improved risk estimates and increased detection of drug event reporting among the various simulated dynamics compared to the PRR. Detection methods that capture temporal adverse drug event dynamics within observational databases can improve our understanding of the interactions between child developmental biology and adverse drug effects.

## Results

### Pediatric FAERS

There were 339,741 pediatric drug event reports in FAERS, which contained 519,555 unique drug-event pairs. We randomly sampled 500 drug-event pairs to be augmented with simulated drug event reporting dynamics, representing our positive control set. We then randomly sampled another 10,000 complementary drug-event pairs where the underlying data was untouched, representing our negative control set. We showed there was no significant difference in the amount of drug-event reporting between FAERS and the negative control (2-sample Student t-test p-value=0.92) or positive control (p-value=0.87) drug-event pairs (Figure 1).

**Fig. 1.**
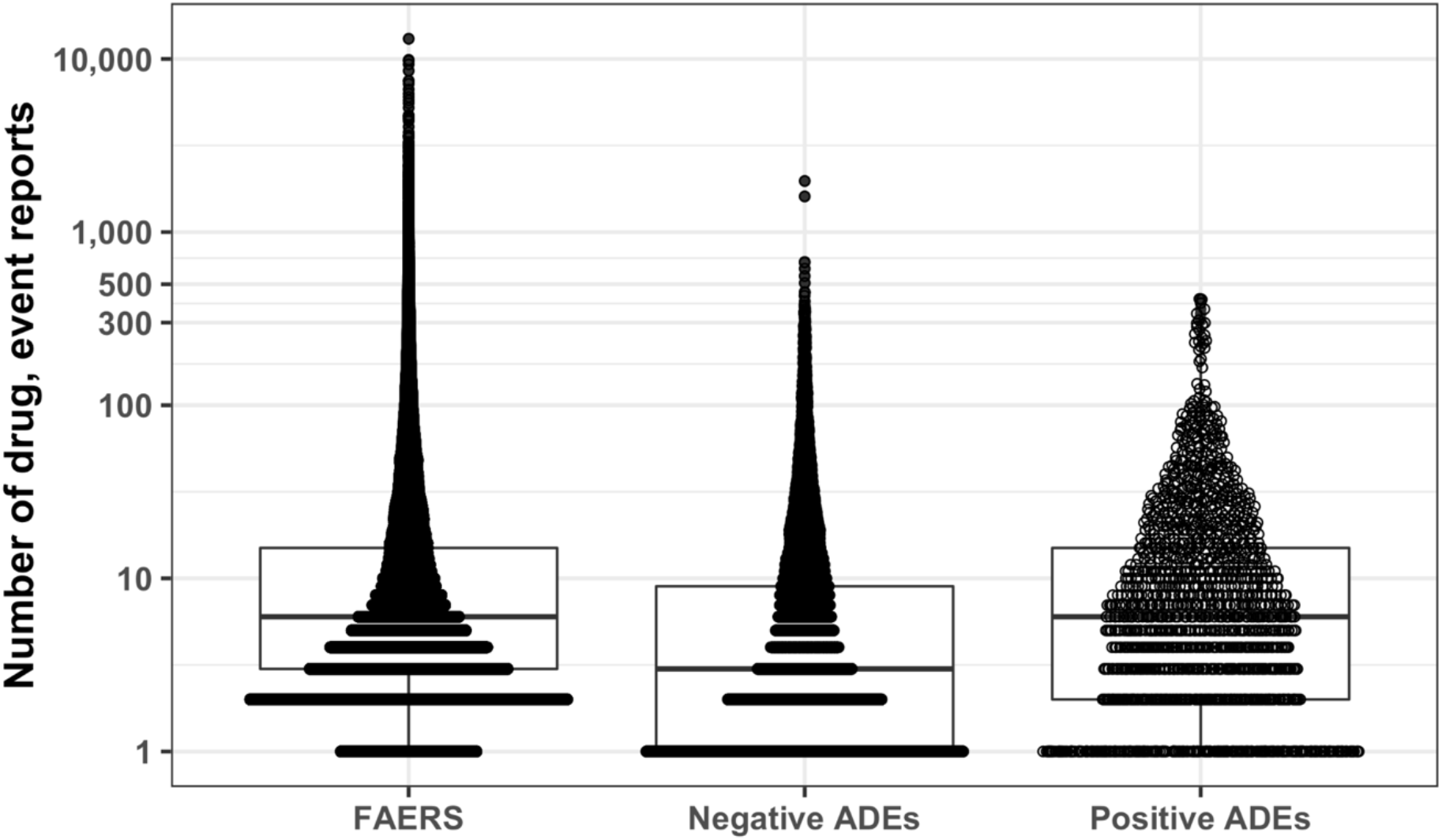
Comparison of drug event reporting in drug-event datasets. A boxplot summary overlayed by the amount of drug event reports for drug-event pairs between (pediatric) FAERS (N=519,555), the positive control set (N=500), and the negative control set (N=10000). ‘N’ is the sample size.

### Data simulation and augmentation

We augmented the 500 drug-event pairs in the positive control set with simulated drug event reporting across child development stages (see Methods). Augmenting the positive control data with drug event reporting dynamics did not have a systematic effect on the amount of drug event reporting compared to the untouched negative control set (Figure S1). However, applying the PRR and GAM detection methods onto the positive control data showed the ADE risk scores reflected the simulated dynamics classes (Figure 2).

**Fig. 2.**
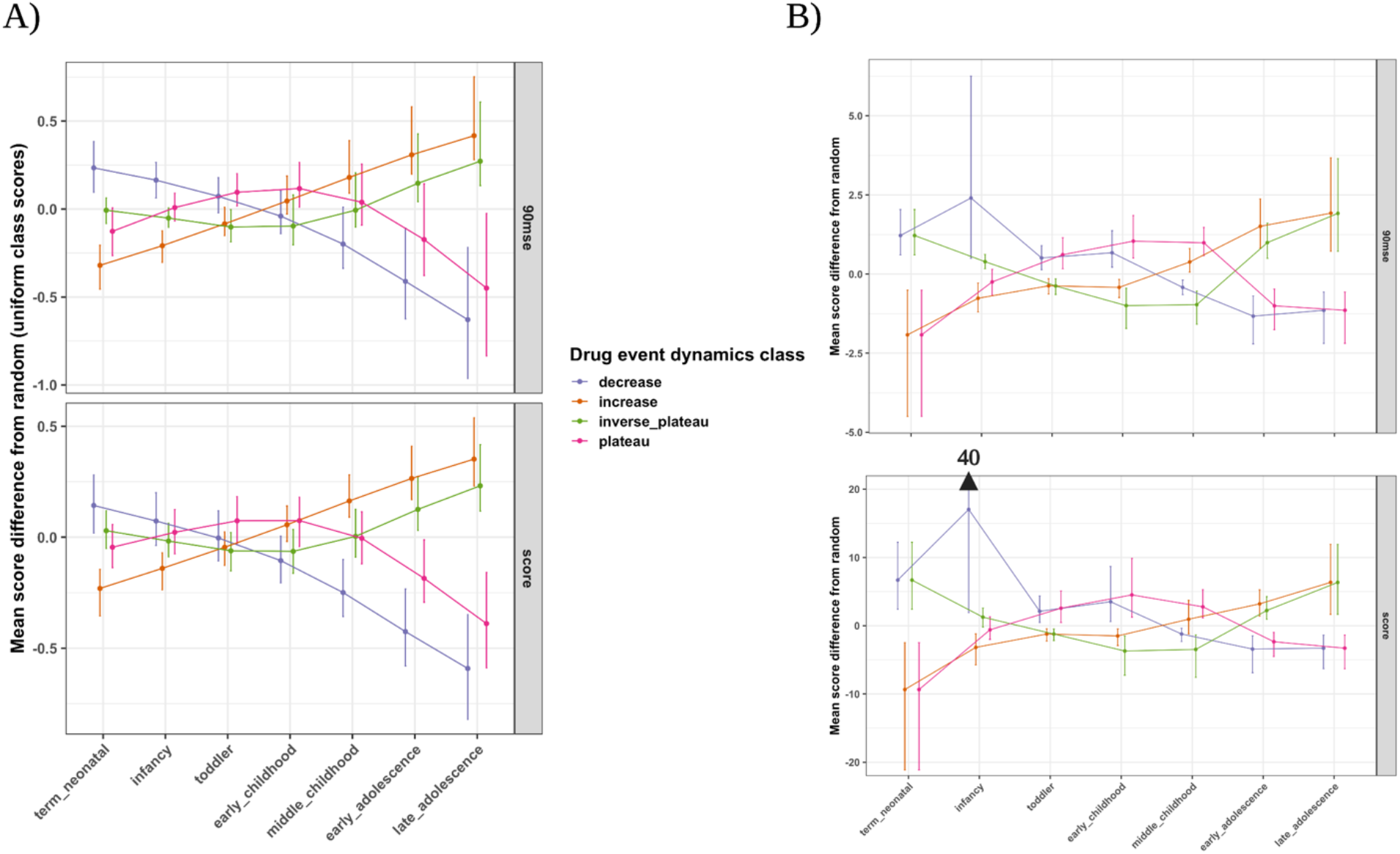
ADE detection method risk score distribution across child development stages. Risk scores resulting from applying the A) GAM and B) PRR ADE detection methods on the positive control drug-event pair data for each dynamics class. The score distributions at each child development stage were produced after 100 bootstraps of the original scores for each method and score type. We show the average difference of the resampled score distributions between a given drug event reporting dynamics class and uniform (random drug event reporting across childhood) with the 95% confidence interval.

The GAM generated ADE risk that resembled normally distributed scores (Shapiro-Wilk test average p-value and 95% confidence interval: 0.45 [0.059, 0.88], 90mse: 0.20 [4.67E-04, 0.92]) in comparison to the PRR (score: 0.11 [1.80E-09, 0.56], 90mse: 0.077 [2.48E-09, 0.93]) at child development stages (Figure 3A). Moreover, 47% of PRR scores were zero and 18% were unable to be computed, on average for drug-event pairs (Figure 3B).

**Fig. 3.**
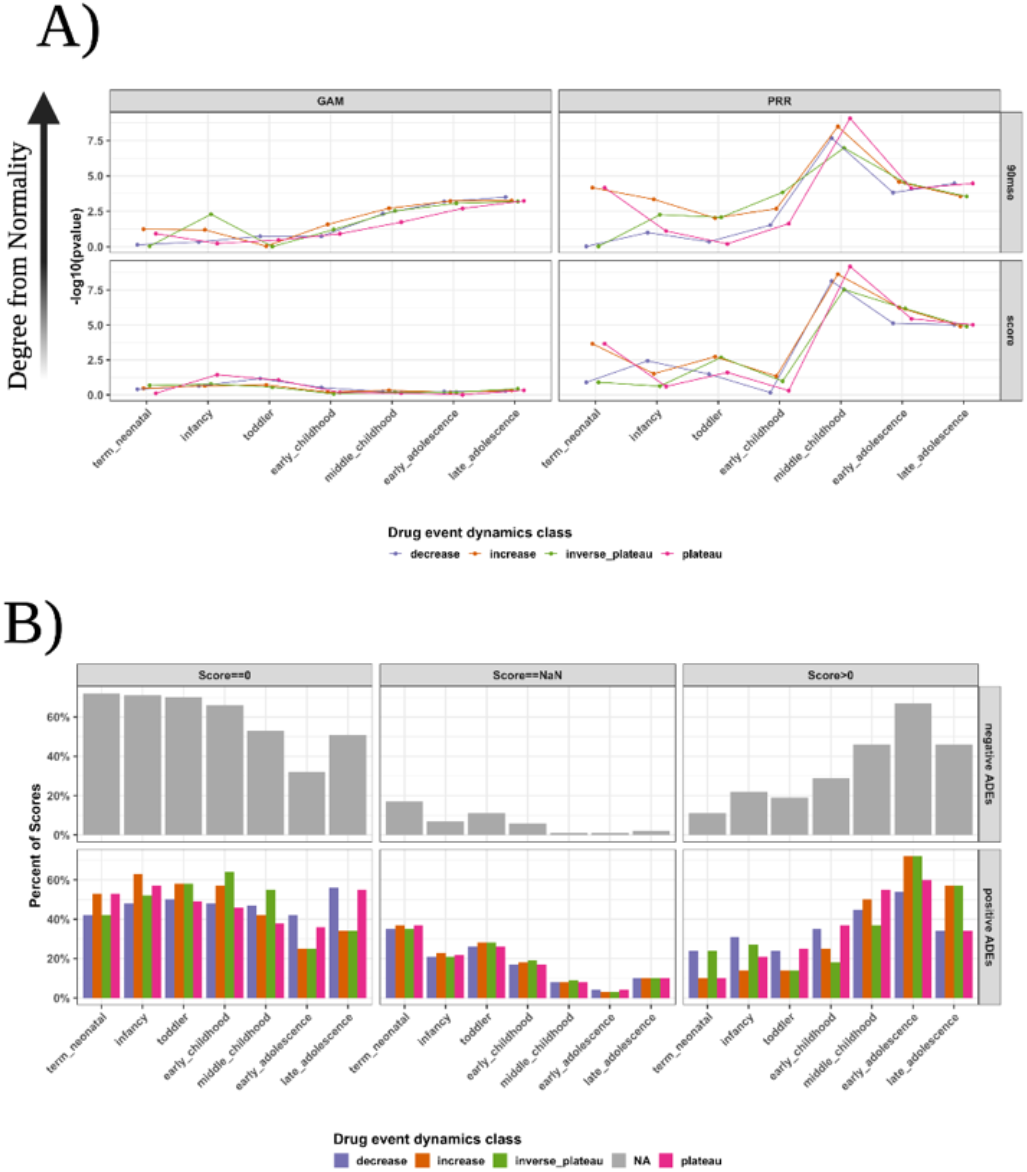
Summary of ADE detection method risk score quality. A) Deviation from a normal score distribution for each method and score type across child development stages. The score distributions were produced after 100 bootstraps of the original scores at each child development stage. The Shapiro-Wilk test calculated a significance probability value for the resampled scores being drawn from a normal distribution. B) PRR detection method risk score quality summary. Across the positive and negative controls as well as drug event reporting dynamics classes, we calculated the number of scores with a zero, NaN (unable to be computed; the drug not reported at a stage or the drug not reported with the event at a stage), or nonzero positive score across child development stages.

### ADE dynamics detection performance

We compared the performance of the GAM and PRR for detecting drug event reporting dynamics (see Methods). Additionally, we further investigated the performance contribution by each child development stage within the dynamics class. We found that the GAM had improved detection of drug event reporting dynamics compared to the PRR both overall (Figure 4A) and within each child development stage (Figure 4B). Moreover, the GAM had similar overall performance (Figure 5) as well as sensitivity (Figure S3) at low drug event reporting compared to the sensitive-by-design PRR.

**Fig. 4.**
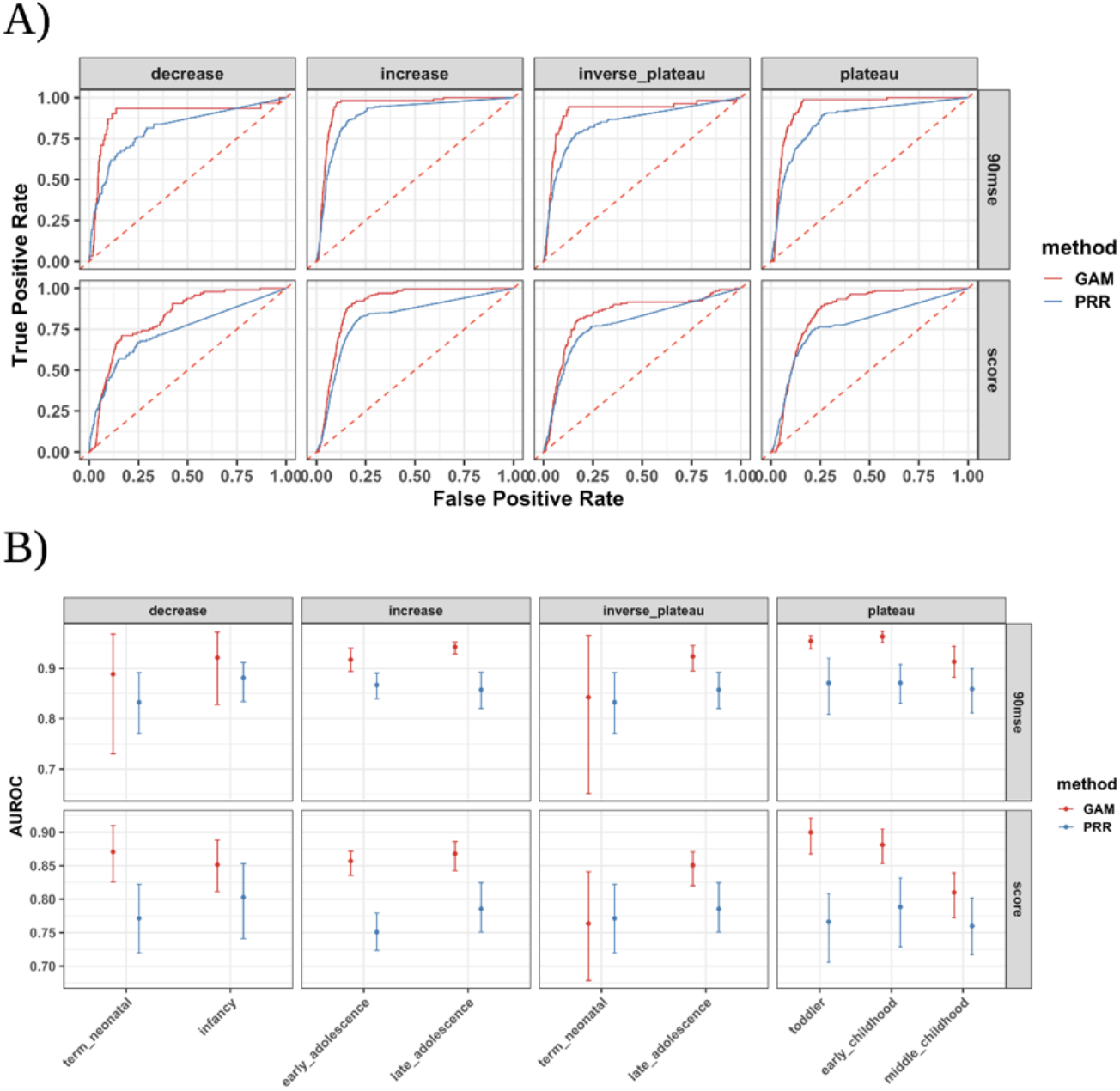
GAM and PRR drug event dynamic detection performance. A) The receiver operating characteristic curves showing the true positive rate versus the false positive rate for each method and score type by drug event reporting dynamics class. B) The area under the receiver operating characteristic curve (AUROC) for each child development stage within each dynamics class.

**Fig. 5.**
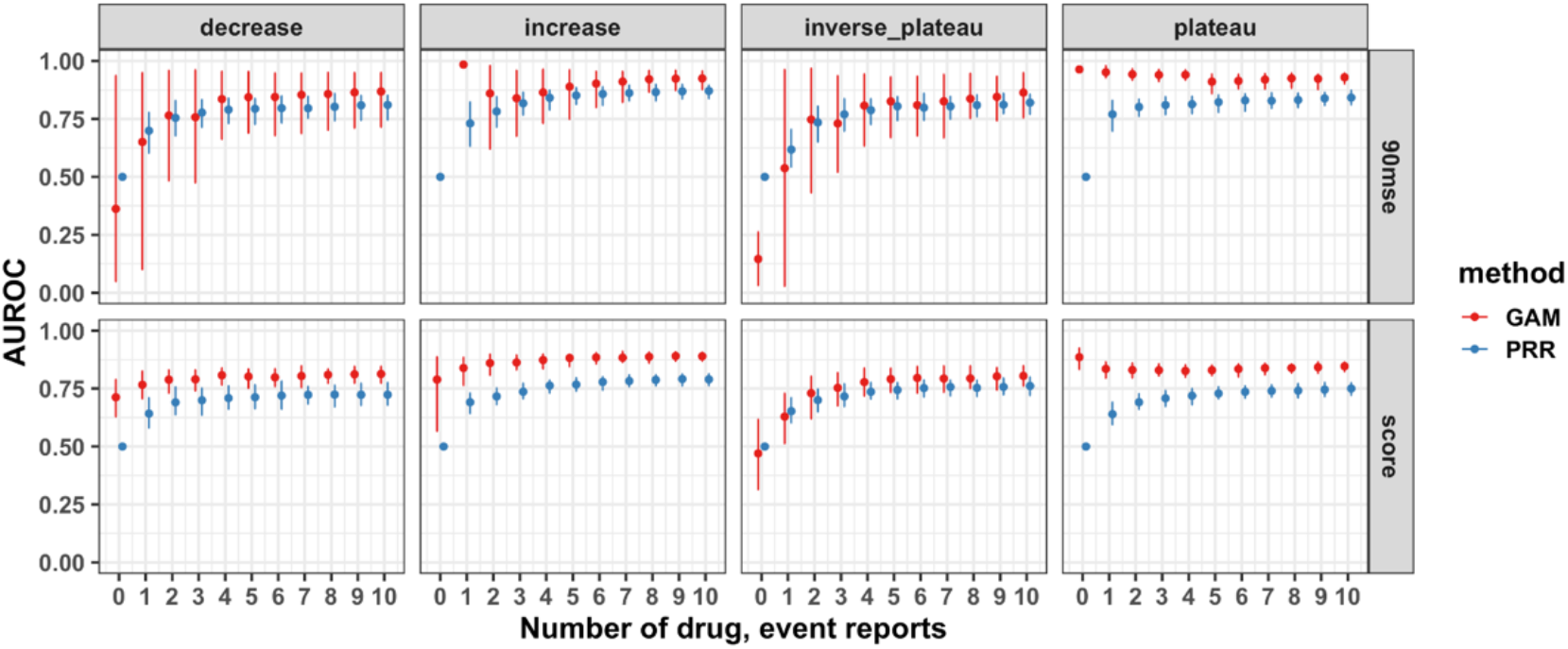
GAM and PRR detection performance at low drug event reporting. The AUROC was computed for each method and score type to detect dynamics only utilizing (drug-event, stage, dynamic) triples with up to a given amount of drug event reports.

### ADE dynamics sensitivity analysis

We investigated the detection of drug event reporting dynamics with increasingly rare adverse events within child development stages (Figure S4 and see Methods). The ADE risk scores generated by the GAM showed dependent, flexible risk estimates across child development stages unlike the PRR (Figure S5). We found that the GAM had significantly higher performance (Figure 6) and sensitivity (Figure S5) to detect the various drug event reporting dynamics as adverse events became rare at child development stages.

**Fig. 6.**
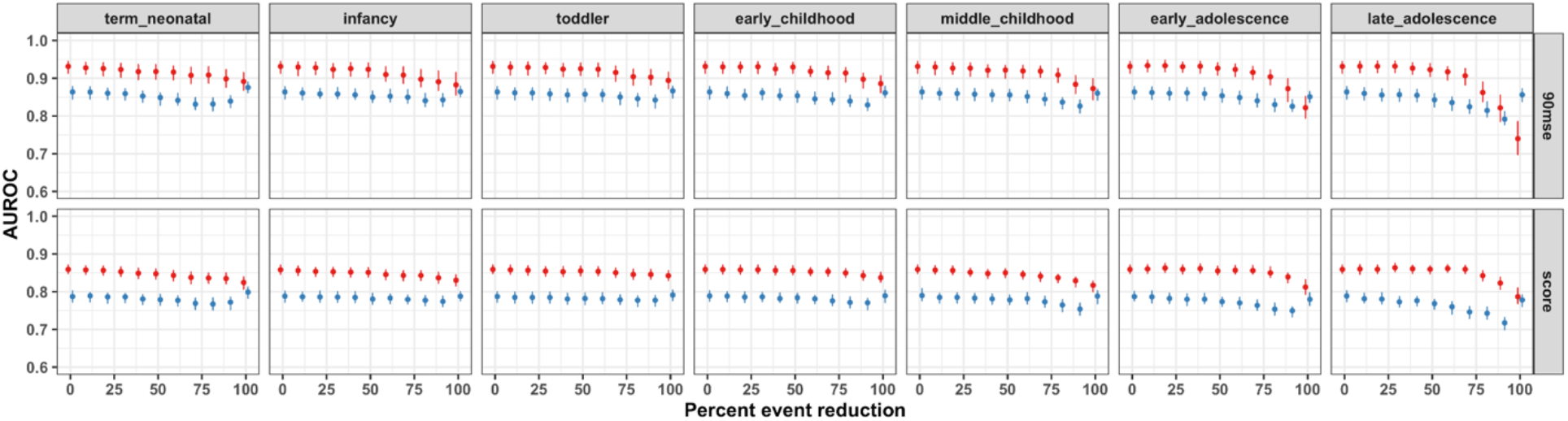
GAM and PRR performance to detect dynamic patterns of rare adverse events. The AUROC for detecting various classes of drug event reporting dynamics as event reporting was reduced at child development stages. The event reporting for drug-event pairs was reduced at 10% decrements only within a specific child development stage. For example, 0% event reporting reduction indicates no reduction in event reporting and 100% event reporting reduction indicates all event reports were removed.

### Real-world validation

We compared the performance of the GAM and PRR for detecting drug-event pairs in a real-world pediatric reference set of 26 drug-event pairs (see Methods and Figure S6). We found that the GAM had slightly improved overall performance and sensitivity compared to the PRR for detecting pediatric adverse drug events (Table 1 and Figure S7). Moreover, we found no difference in the fraction of drug-event pairs with significant ADE risk at child development stages (Table 2; proportion test p-value=0.39). We found that the GAM identified two real-world pediatric drug events with putative dynamic ADE risk (Figure 7). Specifically, the GAM showed periods of lower risk during early and late childhood and higher risk during the middle stages of childhood. While the PRR and GAM performed approximately the same overall, the GAM captured dynamic ADE risk where the PRR did not.

**Table 1.** Real-world pediatric drug-event detection performance. The area under the receiver operating characteristic curve (AUROC) and sensitivity or true positive rate to detect real-world pediatric drug-events observed within FAERS per each method and score type. The prediction threshold for the sensitivity was the null statistic for each method (null threshold: GAM==0; PRR==1). The performance interval is the 95% confidence interval.

|  | Real-world pediatric drug-event detection performance |  |  |  |  |  |  |  |
| --- | --- | --- | --- | --- | --- | --- | --- | --- |
|  | AUROC |  |  |  | Sensitivity |  |  |  |
|  | 90mse |  | score |  | 90mse |  | score |  |
|  | PRR | GAM | PRR | GAM | PRR | GAM | PRR | GAM |
| Performance value | 0.62<br>[0.58, 0.66] | 0.65<br>[0.59, 0.69] | 0.62<br>[0.58, 0.67] | 0.73<br>[0.69, 0.77] | 0.28<br>[0.22, 0.36] | 0.28<br>[0.23, 0.35] | 0.49<br>[0.40, 0.56] | 0.85<br>[0.81, 0.89] |

**Table 2.**
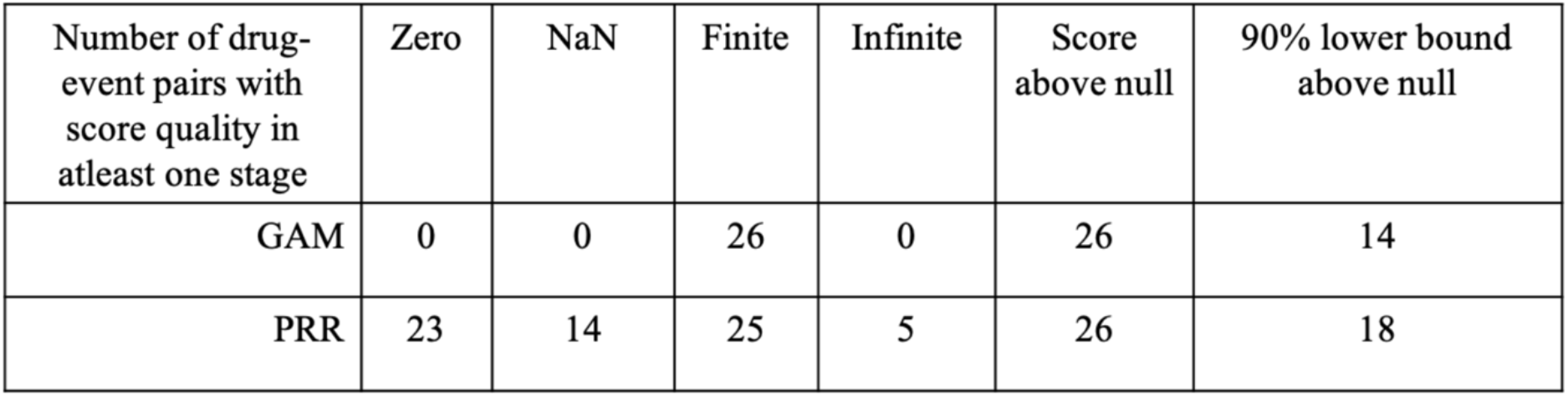
ADE detection method risk score quality and significance on real-world pediatric drug-events. The number of drug-event pairs that contained a child development stage with a risk score of each score quality (null threshold: GAM==0; PRR==1). A 90% lower bound score above the null threshold indicates a significant risk.

| Number of drug-event pairs with score quality in at least one stage | Zero | NaN | Finite | Infinite | Score above null | 90% lower bound above null |
| --- | --- | --- | --- | --- | --- | --- |
| GAM | 0 | 0 | 26 | 0 | 26 | 14 |
| PRR | 23 | 14 | 25 | 5 | 26 | 18 |

**Fig. 7.**
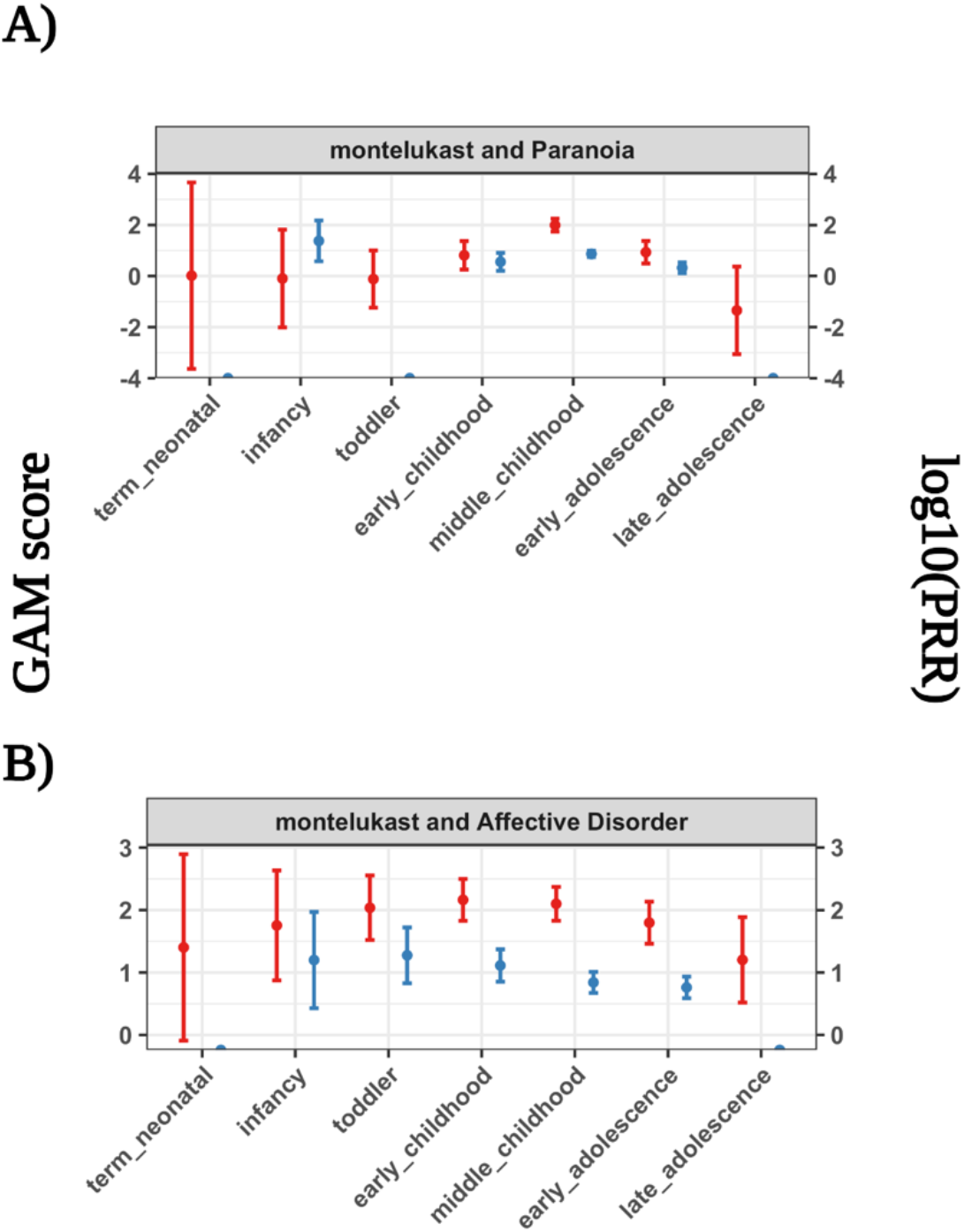
GAM and PRR detection scores on putative real-world dynamic adverse drug events. We highlight two psychiatric adverse events, A) Paranoia and B) Affective Disorder, from exposure to the drug montelukast exhibiting dynamic ADE risks across child development stages.

## Discussion

Children undergo a period of dynamic growth and development, presenting a challenge in identifying and evaluating adverse drug events^10,36^. We hypothesize that dynamic ontogenic processes as children grow and develop may be reflected by temporal drug event reporting in the population. We presented the first study to evaluate drug event reporting patterns across childhood in large observational data. We found that GAMs, a population modeling technique, outperformed the PRR, a population stratification method, as well as generated robust risk scores to detect adverse drug events during childhood. This work represents a first step in transitioning from performing event surveillance towards uncovering putative mechanisms of pediatric adverse drug events.

The goal of our study is to improve the specificity of top-down data mining for generating pediatric drug safety hypotheses. Our study hypothesis was temporal drug event reporting trends found in observational data are dependent on ontogeny, which exhibits high and low molecular and physiological levels throughout childhood^7,37,38^. To test this within a top-down approach, we generated temporal trends in observational data to correspond with temporal trends from ontogeny as opposed to identifying temporal trends from frequency^39^ or feature-derived^40,41^ measures directly from observed data. This motivated both simulating dynamic drug event reporting rates and then augmenting real-world data to generate different classes of dynamic drug event reporting trends. While we simulated dynamic drug event reporting rates, we showed that augmenting the FAERS data did not change the overall characteristics of the pediatric drug reports. This was crucial for establishing the use of real-world drug event data to evaluate hidden dynamic reporting trends. Importantly the ADE detection methods were in fact able to identify the simulated dynamics within the data. The data simulation and augmentation of FAERS laid the foundation for evaluating statistical methods to investigate ontogenic-mediated adverse event mechanisms.

We found that the generalized additive model (GAM) showed improved detection of dynamic drug event reporting compared to the proportional reporting ratio (PRR). While the PRR produces ADE risk scores that were more erratic and unable to be computed, the GAM scores were both more flexible and robust. The GAM assumes a flexible relationship yet reduces ‘wiggliness’ to stable risk estimates based on observed data^42,43^. While bayesian modeling techniques such as Monte Carlo Markov Chain can also learn flexible relationships from observed data, these models still require expert knowledge to build, implement, and interpret^44^. The GAM, on the other hand, generates an interpretable smooth relationship in a familiar regression framework^32^ that shares information across child development stages. Using this shared information framework, the GAM was able to detect injected dynamic ADE risks across childhood even when drug event reporting was low. We further showed that the GAM not only generated visually dynamic ADE risk when injecting dynamics, but we also identified putative dynamic risk for real-world psychiatric adverse events from exposure to montelukast medication (Figure S8). We demonstrated that GAMs can be used to detect dynamic reporting of adverse drug events by sharing information across child development stages.

This study has some limitations. First, observational data has inherent bias and confounding factors which may affect both the sample of drug-event pairs in our study as well as the performance of the detection methods. We showed that the random sample of drug events correspond to the reporting patterns found in the FAERS database. Also, performing a power analysis allowed for identifying drug events for which the detection methods were able to identify the dynamic reporting to provide a fair performance comparison. Second, other regulatory agencies, such as the Food and Drug Administration and European Medicines Agency, define pediatric age ranges for development stages by different methods. While varying child stage definitions were not explored here, we chose stages defined by NICHD that were established after consultation and agreement among several US-based organizations such as the American Academy of Pediatrics and the Centers for Disease Control and Prevention^45^. Third, fixed development stages may serve more useful in drug regulations and trial design than representing dynamic child growth and development. Nevertheless, the detection performance and risk scores for both methods could only be compared when considering data found within child development strata. Fortunately, the further advantage of the GAM is its ability to model childhood as a continuous period using age without restrictive strata. This increases the sharing of information for identifying adverse event risk during childhood which may cross development stages and affect specific periods during childhood.

## Conclusion

In this study, we evaluated ADE risk detection methods to identify dynamic drug event reporting within observational data. By simulating drug event reporting and augmenting simulated rates into existing observational data, we can make comparisons between methods to detect dynamic drug event reporting patterns. We found GAMs result in more robust scores, overall improved performance to detect dynamics, and improved ability to detect simulated and real-world pediatric drug-events compared to the state-of-the-art PRR method. This study lays the foundation to detect and evaluate pediatric adverse drug events for ontogenic-mediated mechanisms.

## Data Availability

The datasets and code supporting the conclusions of this article are available in the evaluating_ontogenic_ade_risk Github repository, DOI: 10.5281/zenodo.4585585.

https://github.com/ngiangre/evaluating_ontogenic_ade_risk

## Methods

### ADE data source

We retrieved drug event reports from the Food and Drug Administration’s openFDA^46^ download page, utilizing an API key with extended permissions, containing the FAERS data. Using custom python notebooks and scripts available in the ‘openFDA_drug_event-parsing’ github repository (DOI: 10.5281/zenodo.4464544), we extracted and formatted all drug event reports prior to the third quarter of 2019. Data fields included the safety report identifier, age value, age code e.g. year, adverse event MedDRA concept code (preferred terms), and drug RxNorm code (various) used in our analyses. The age value was standardized to year units for categorizing reports into the 7 child development stages according to the Eunice Kennedy Shriver National Institute of Child and Human Development^45^. Adverse drug event MedDRA codes were mapped to standard concept identifiers using concept tables^48^ from the OMOP common data model. The drug RxNorm code was similarly translated to the standard RxNorm concept identifier (ingredient level) in OMOP and was further mapped to the equivalent ATC concept identifier (ATC 5^th^ level) using the concept relationship table. The occurrence of an adverse drug event is defined as any safety report where both the adverse event and drug concepts are reported together. The pediatric report space for any adverse drug event is all reports which have age above zero and less than or equal to 21 years old which is the upper bound for the late adolescence child development stage. The drug event data for a given drug-event pair composed of 339,741 safety reports with a binary indicator for reports of the event and drug, as well as the category of NICHD child development stage for the report’s patient.

### Simulated ADE dynamics

The objective of this study was to evaluate detection of drug-event reporting as the reporting rate changes across child development stages with varying dynamics and effect sizes. We assert that reporting dynamics during childhood reflect ontogenic profiles observed on molecular, functional, and structural levels^7,37,38^.

We simulated dynamic ADE reporting by combining hyperbolic tangent functions that produced symmetric probability distributions around a given effect size to define the probability of event reporting at drug reports. These dynamic reporting classes represent nonlinear trends of drug-event reports across childhood. The average drug and event reporting across reports equaled the event reporting rate multiplied by a fold change factor resulting in the effect size of dynamic drug event reporting. The fold change followed a negative exponential distribution with rate parameter 0.75 resulting in a fold change distribution ranging from 1 to 10 (Figure S9). The simulated reporting probabilities were distributed to safety reports in age ascending order reflecting a desired dynamic distribution of ADE reporting across childhood. We designed 5 different dynamic reporting rates, namely ‘uniform’ (random), ‘increase’, ‘decrease’, ‘plateau’, and ‘inverse_plateau’ (Figure S10).

### ADE data augmentation

We augmented the original drug event data from FAERS with the simulated drug event reporting dynamics. We randomly selected 500 drug-event pairs to be the positive control set. We augmented the drug event data for each pair with dynamics previously described that we want to detect. We then randomly selected 10,000 mutually exclusive drug-event pairs to be the negative control set which were not augmented and represented reporting of drugs with events within FAERS. Differences of the average drug event reporting between the drug-event sets was computed by comparing 10 million resamples of each distribution.

Augmenting the positive control drug-event pairs resulted in 5 sets of 500 drug-event pairs, forming (drug-event, stage, dynamic) triples. The (drug-event, stage, uniform dynamic) triple scores were the reference distribution for comparing the average difference in scores, after 20 resamples, with ADE risk scores from the other dynamics classes.

### ADE detection methods

We applied two ADE detection methods to the positive and negative control drug-event sets. We chose a population stratification (PRR) and modeling (GAM) method to evaluate detection of ADE dynamics when stratifying the data or by sharing information across child development stages, respectively.

We employed the Proportional Reporting Ratio (PRR):

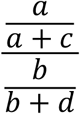

where ‘a’ is the number of reports with the drug and event, ‘b’ is the number of reports without the drug and with the event, ‘c’ is the number of reports with the drug and without the event, and ‘d’ is the number of reports without the drug or event of interest. The resulting score is the event reporting prevalence with the drug compared to without the drug. We generated PRR scores for each child development stage resulting in 7 scores for each drug-event pair. The PRR scores were log10 transformed when conducting the Shapiro-Wilk test for normality.

We also evaluated the logistic generalized additive model^49^ (GAMs):

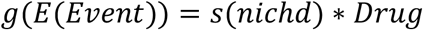

where *g* is a logit link function, *E(Event)*is the expected value of event reporting, *s* is a spline function with a penalized cubic basis, *nichd* is the child development stage of the report’s subject, and *Drug* is an indicator i.e. 0 or 1 of drug reporting. Details for GAMs can be found at references^42,50^ and we specified the model using the *mgcv* package in R.

Briefly, the GAM is a flexible statistical model that captures nonlinear effects of covariates onto a response. In this paper, we model the effect of the child development stage interacting with drug reporting on the reporting of an event where the event is the reporting of the MedDRA preferred term and the drug is the reporting of the ATC 5^th^ level drug concept. The *s*() function is a spline function where the interaction of the child development stage (main effect) and the drug (interaction using the ‘by’ variable) is modeled according to a set of basis functions. Each development stage defines the knot (7 in total) in which the expectation of event reporting is quantified. In the spline function, a penalized cubic spline basis (bs=‘cs’) is used for fitting the basis functions where the first and second derivative of the event expectation is zero at each knot, resulting in a smooth event expectation across stages. To mitigate overfitting or ‘wiggliness’, we used a penalized iterative restricted likelihood approach, called ‘fREML’, with a wiggliness penalty in the objective function. Fitting the GAM model (using the ‘bam’ function and discrete=T) produces coefficient terms, similar to beta coefficients in logistic regression, for each child development stage for the association of the adverse event being reported in interaction with reporting the drug. We generated GAM scores for each child development stage resulting in 7 scores for each drug-event pair. It is important to note that all GAM scores produced were finite, nonzero values.

The scores generated by each method have different variations and uncertainty in the estimated population value. We additionally determined the lower confidence bound in which the population-based score would be greater than 90% of score replicates. The population score and the 90% lower confidence bound, called ‘score’ and ‘90mse’ respectively, are the score types for each method.

### ADE dynamic detection power analysis

We performed a power analysis to determine which of the positive control drug-event pairs could be detected for each method and score type. The generated scores may not show a drug and event association (score above the null statistic or a significance association) for a child development stage due to the method’s different assumptions and biases when applied onto observational data. To mitigate these issues, we determined the drug event data characteristics, namely the number of drug reports and the effect size, for each method in which reporting dynamics could be detected at or above *t* = 80% power or true positive rate. Specifically, for the (drug-event, stage, dynamic) triple scores in the positive control set, we determined the power to differentiate scores at high reporting rates about a given score threshold (GAM score threshold==0; PRR score threshold==1). The reporting rates were higher at different child development stages for each dynamics class e.g. the ‘increase’ dynamics class had higher reporting at the ‘early_adolescence’ and ‘late_adolescence’ stages (Table S1). The scores from (drug-event, stage, dynamic) triples with a high reporting rate were only considered for reflecting dynamic drug event reporting associations. The scores from (drug-event, stage, dynamic) triples with a low reporting rate were not considered further due to spurious scores generated at stages without injected signal. The drug event characteristics were determined for both the estimated population score (‘score’) and the 90% lower bound score (‘90mse’) that represent scores with lower and higher confidence, respectively, for the ‘true’ population score.

Choosing drug-event pairs at or exceeding the characteristics for each method and score type at or above *t* = 80% power resulted in a superset of (drug-event, stage, dynamic) triples designated as positives in a reference standard for each drug event reporting dynamics class (Table S2). The negative control set contained the same (drug-event, stage) doubles or 70,000 scores for each reference standard. Excluding the drug-event scores generated by the uniform class, there were 4 reference standards of positive and negative drug-event pairs for each ADE reporting dynamics class used for detection performance evaluation.

### ADE dynamic detection performance

We evaluated the GAM and PRR methods to detect drug event reporting dynamics across the child development stages. Specifically, we determined the performance in differentiating scores from (drug-event, stage, dynamic) triples in the positive control set versus the negative control (drug-event, stage) score doubles. The positive control set contained a superset of the 500 (drug-event, stage, dynamic) score triples (Table S2). The negative control set contained the same (drug-event, stage) doubles or 70,000 scores for each reference standard. For each of the four reference standards, we quantified performance metrics including the sensitivity, specificity, and area under the receiver operating characteristic (AUROC) curve using the R package *ROCR* for each detection method and score type. Confidence intervals for the AUROC were calculated through bootstrapping (100 resamples) the score distributions and calculating performance metric values.

### Dynamics sensitivity analysis

We assessed the sensitivity of the ADE detection methods to detect drug event reporting dynamics within child development stages. We artificially reduced, at 10% decrements, the event reporting rate at each child development stage separately. Specifically, at each reduced stage, we determined the sensitivity of each method and score type to detect (drug-event, stage, dynamic) score triples compared to the same negative control (drug-event, stage) score doubles at that same reduced stage. Sensitivity was assessed iteratively at the 10% decrements within each child development stage. We calculated the AUROC and power metrics to quantify sensitivity to drug event reporting dynamics for each method and score type.

### Real-world ADE validation

We applied the ADE detection methods on observed FAERS data for drug-event pairs within the pediatric drug-event reference standard from the Global Research in Pediatrics consortium^51^. A machine-readable dataset can be found at the ‘GRiP_pediatric_ADE-reference_set’ github repository (DOI: 10.5281/zenodo.4453379). We assigned drug-event pairs with epidemiological or mechanistic evidence in children (Control==‘C’) as the positive class (N=26), and the cross-product of all drugs and events that were complementary to drug-event pairs in the reference set as the negative class (N=123). We calculated the AUROC using the *ROCR* package in R and the true positive rate using the null statistic of each method as the prediction threshold.

## List of Abbreviations

ADE: adverse drug event
FAERS: Food and Drug Administration Adverse Event Reporting System
GAM: generalized additive model
PRR: proportional reporting ratio
NICHD: national institute of child and human development
AUROC: area under the receiver operating characteristic curve
TPR: true positive rate
ATC: anatomical therapeutic class
DOI: digital object identifier

## Availability of Data and Materials

The datasets and code supporting the conclusions of this article are available in the ‘evaluating_ontogenic_ade_risk’ Github repository, DOI: 10.5281/zenodo.4585585.

## Declarations

None.

---

**Fig. S1.**
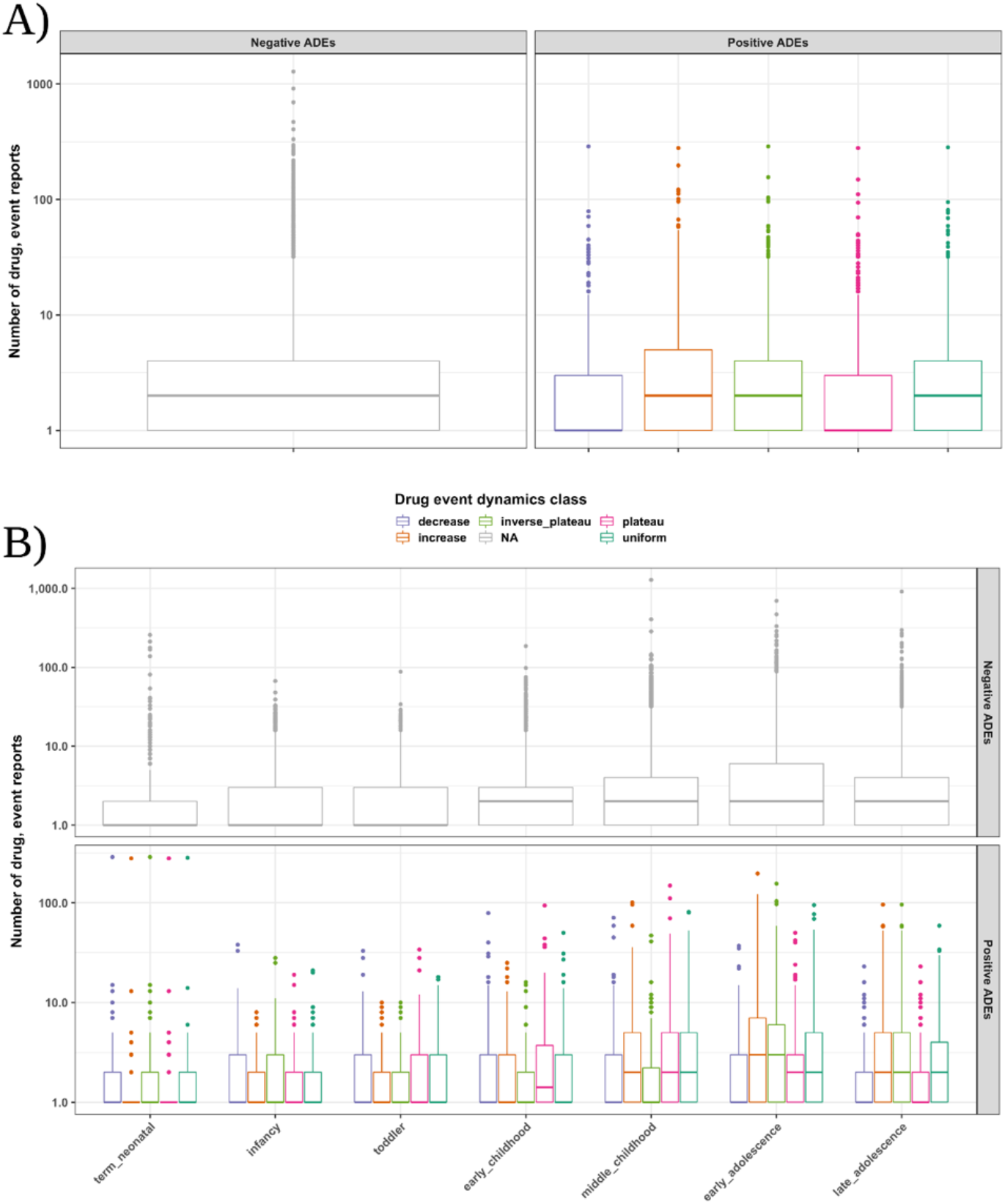
Drug event reporting for positive and negative controls. The distribution of drug-event reporting between positive and negative control set drug-event pairs across A) dynamics classes and B) child development stages.

**Fig. S2.**
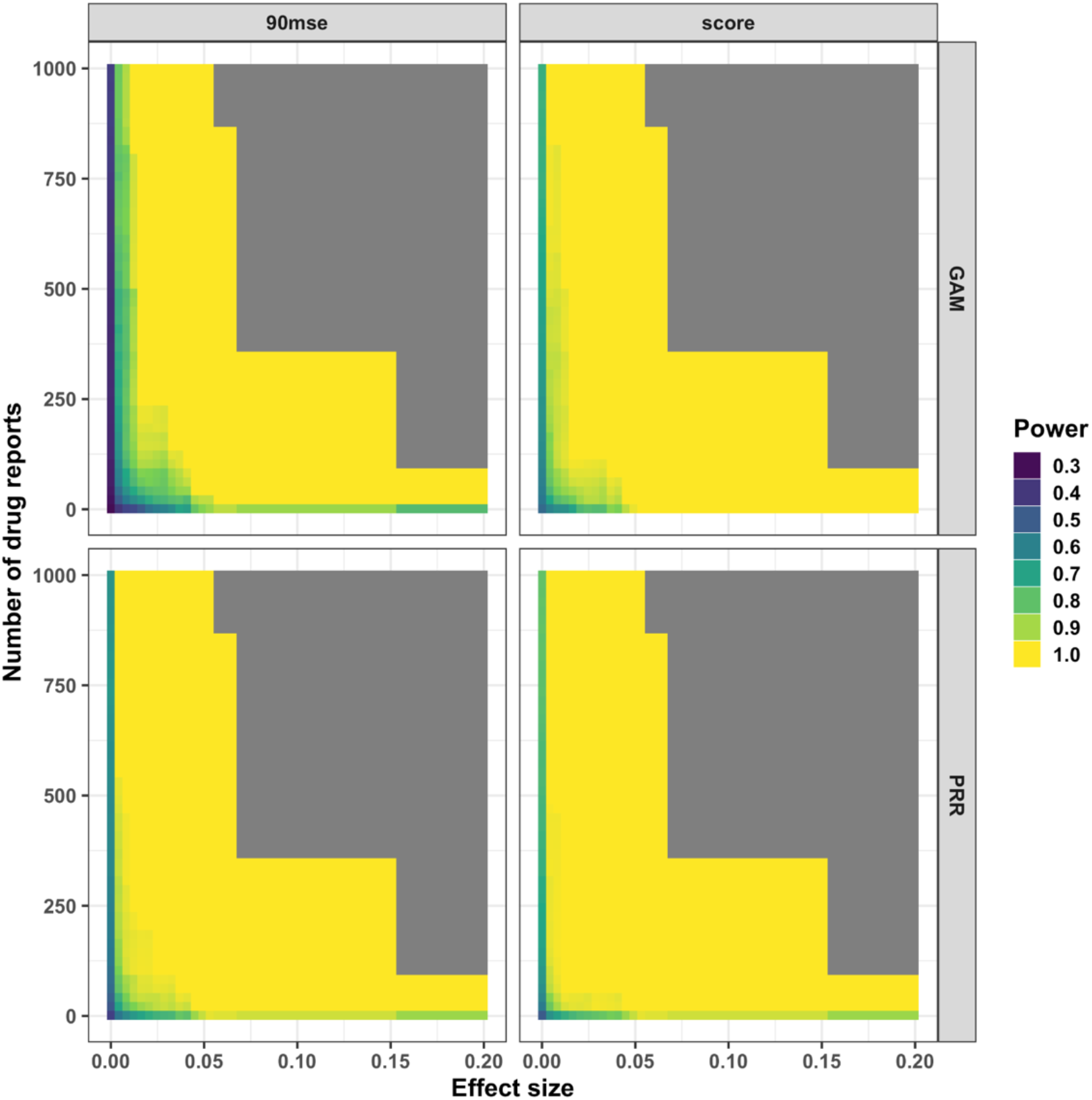
Power graph detecting drug-event pairs with dynamics for each method and score type. The number of drug reports and effect size determined the scores evaluated to detect an association between the drug and event co-occurrence. The scores used were from child development stages with a high reporting rate determined by the dynamic class.

**Fig. S3.**
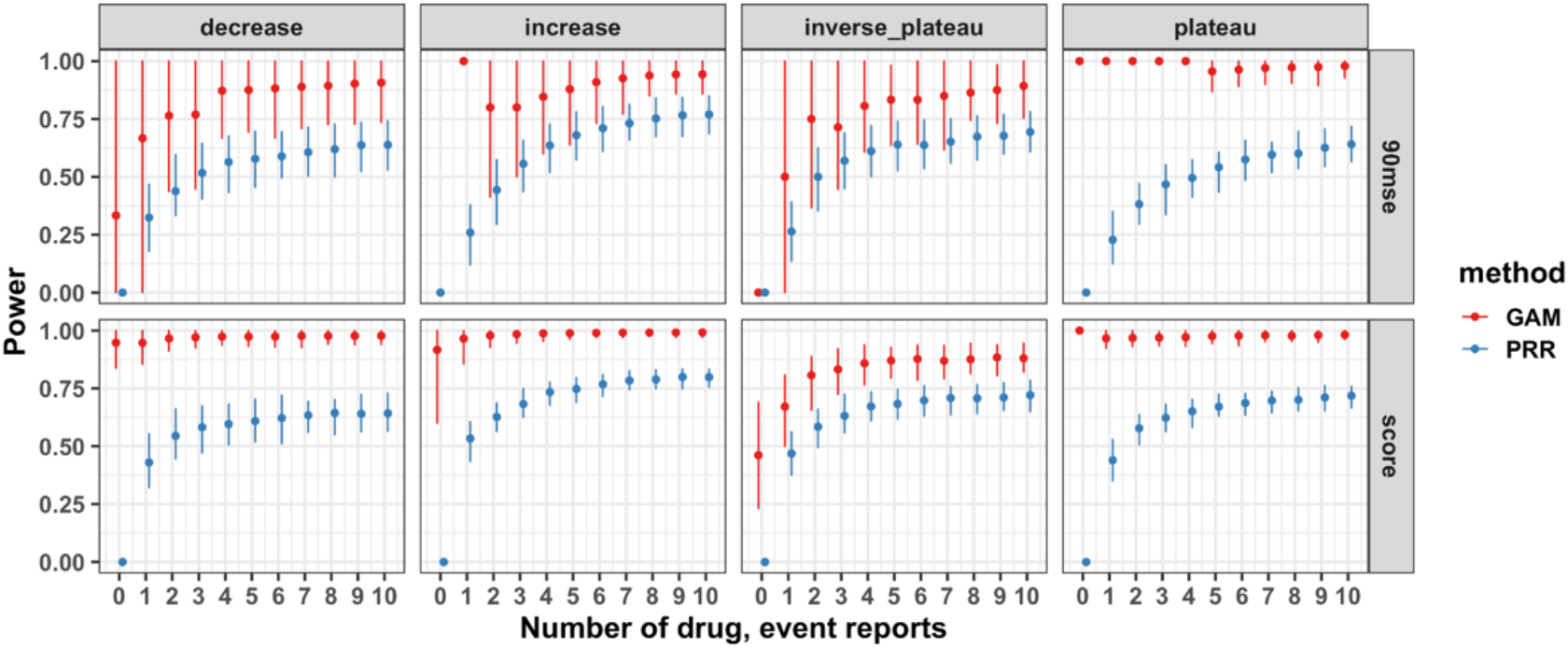
GAM and PRR detection sensitivity at low drug event reporting. The sensitivity or true positive rate was computed for each method and score type to detect dynamics only utilizing (drug-event, stage, dynamic) triples with a given amount of drug event reports.

**Fig. S4.**
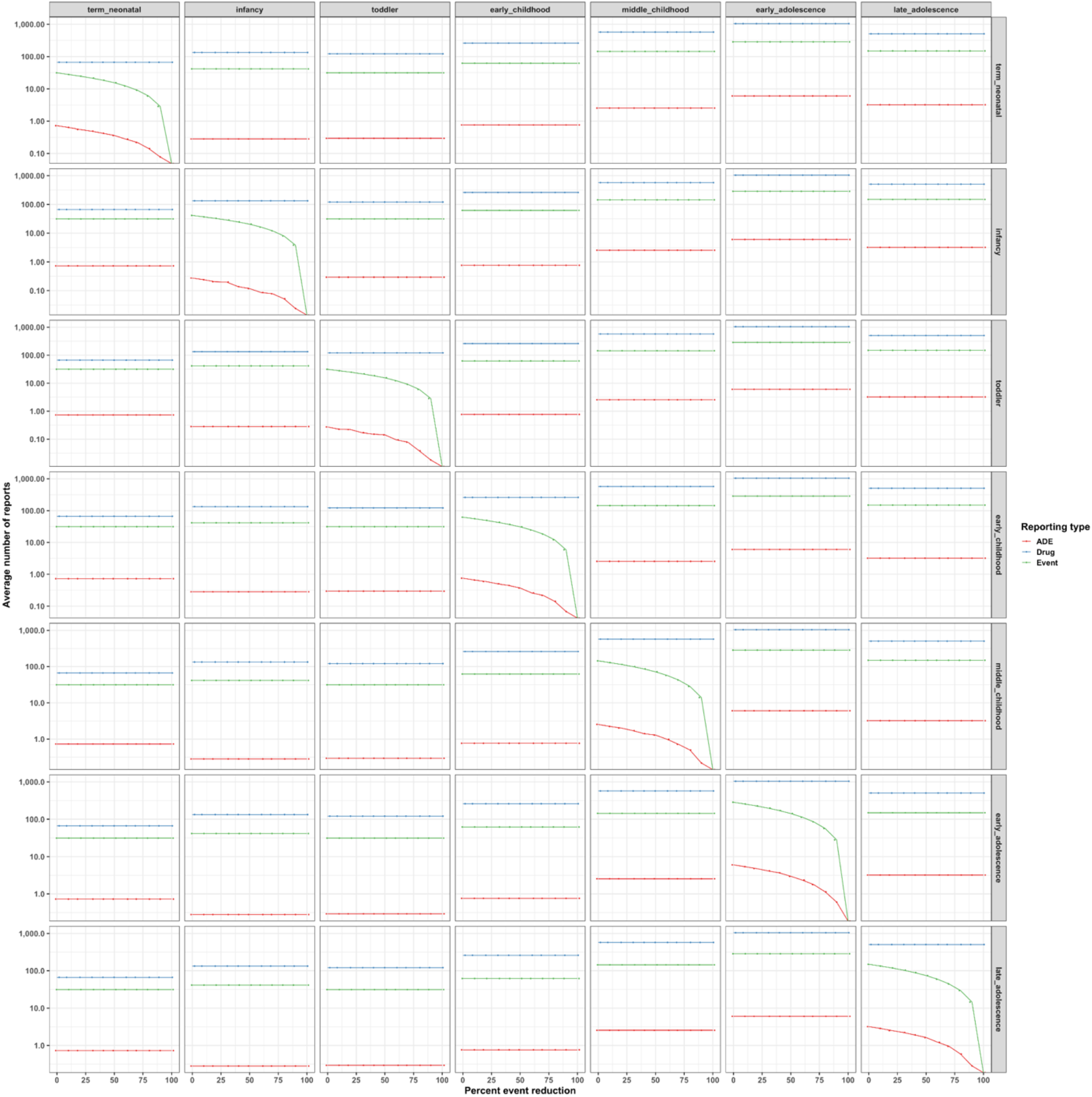
Drug and event reporting as adverse events become rare at child development stages. The average number of drug, event, and drug event reports are shown at each child development stage as event reporting is reduced by 10% at each child development stage.

**Fig. S5.**
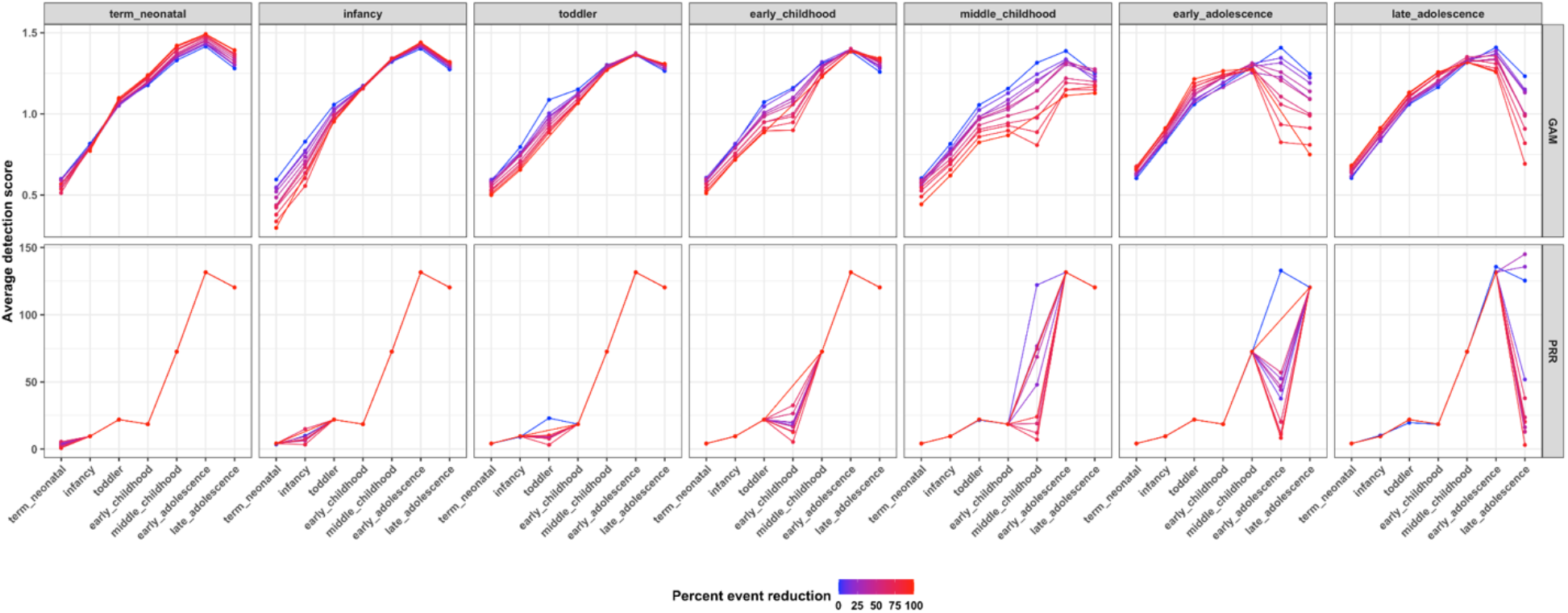
GAM and PRR detection scores across childhood as adverse events become rare. The average ADE risk scores are shown at each child development stage as event reporting is reduced by 10% at each child development stage.

**Fig. S6.**
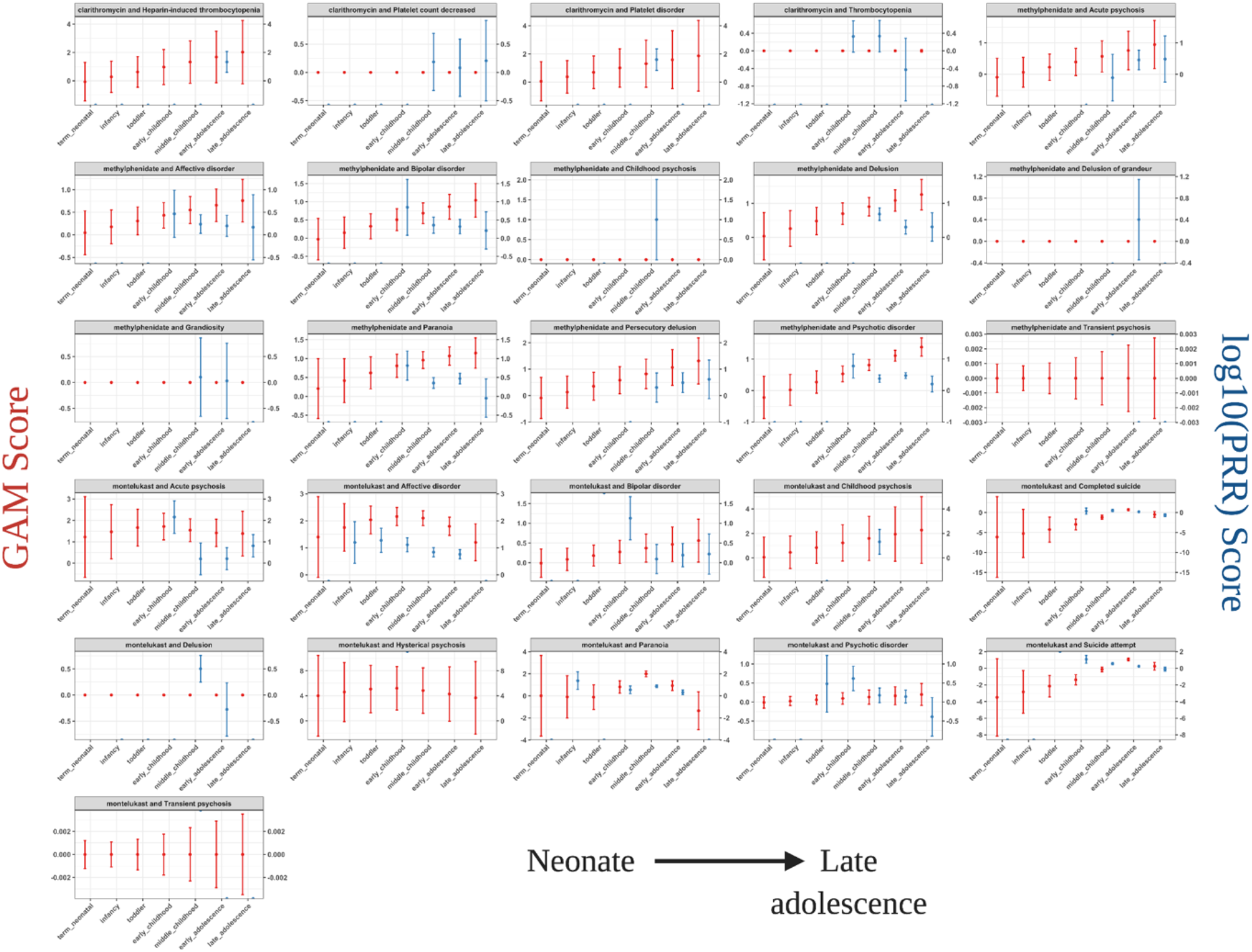
GAM and PRR score comparison for real-world pediatric drug-events. Application of each ADE detection method on drug-event pairs from the clinically-validated GRiP pediatric reference set. The population score and 90% confidence interval are shown for each detection method. Only drug-event pairs with epidemiological or mechanistic evidence in children are shown.

**Fig. S7.**
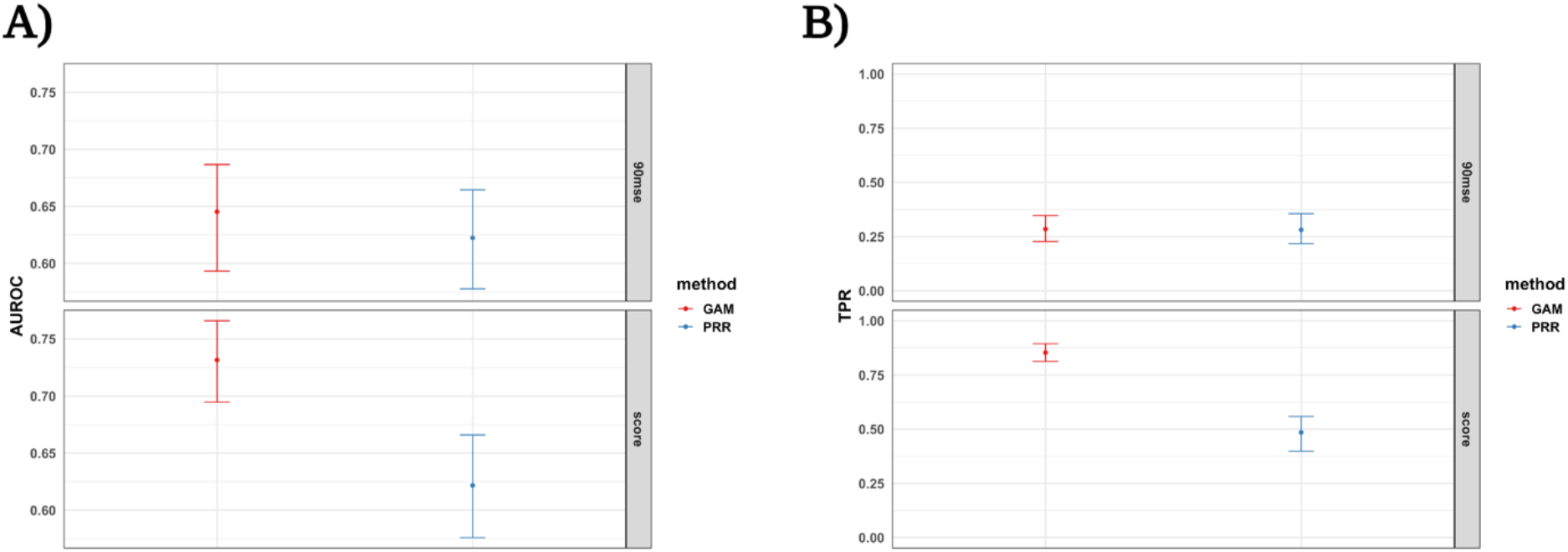
GAM and PRR drug event real-world drug-event detection performance. A) The area under the receiver operating characteristic curve (AUROC) for detecting risk in real-world pediatric drug-event pairs. B) The true positive rate (TPR) for detecting risk in real-world pediatric drug-event pairs. Intervals represent the 95% confidence interval for the performance metric.

**Fig. S8.**
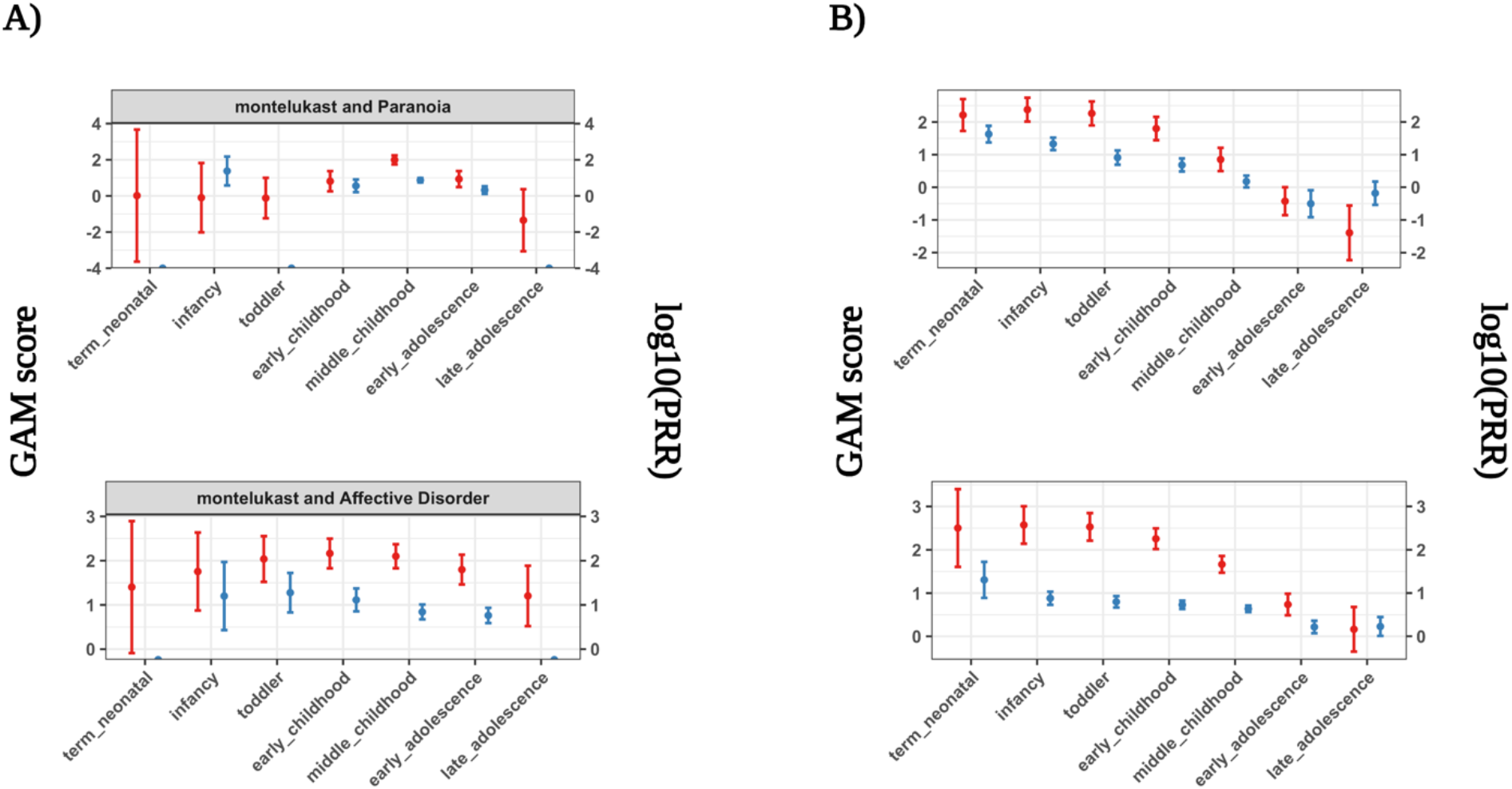
GAM and PRR detection scores on sample real-world and simulated drug event data. We highlight two drug-event pairs from the A) real-world and B) simulated positive control set where scores across child development stages reflect dynamic ADE risks.

**Fig. S9.**
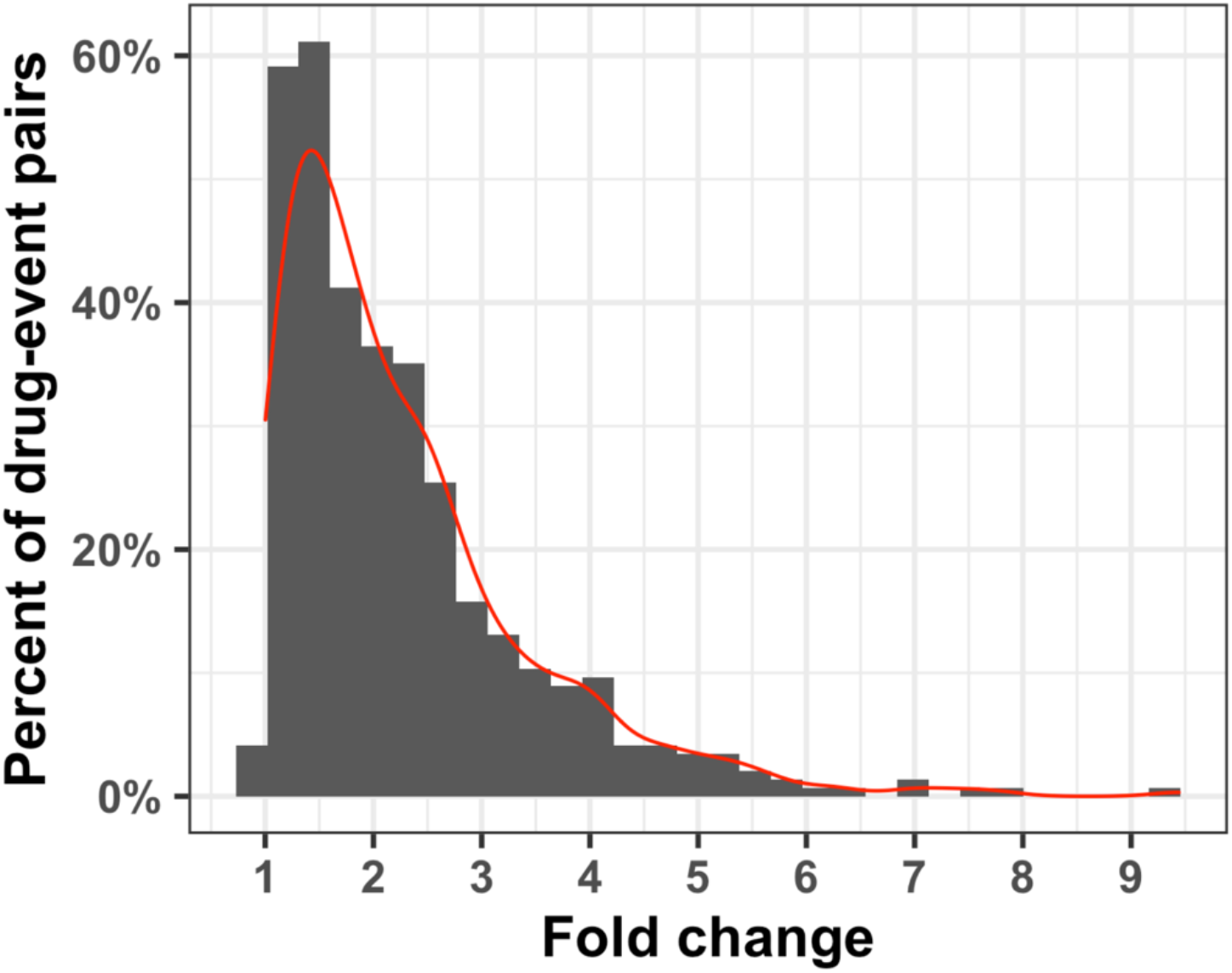
Distribution of fold-changes for generating drug event dynamics of varying magnitudes. Distribution of fold-changes randomly assigned to drug-event pairs in the positive control set for each dynamics class.

**Fig. S10.**
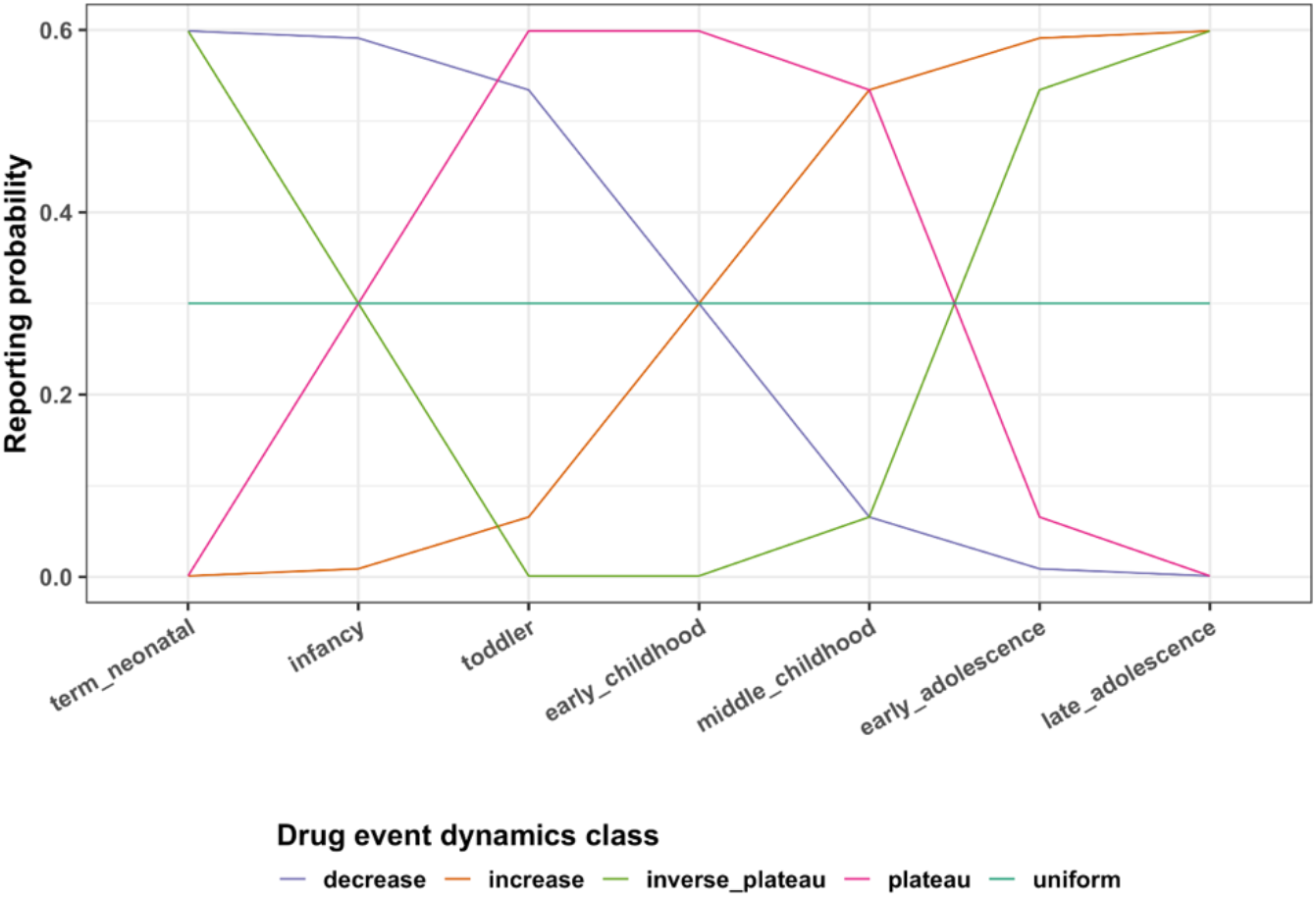
Drug event reporting probability rates across child development stages. Each dynamics class represents a drug event reporting probability trend for drug-event pairs in the positive control set. An effect size, here 0.3, determined the magnitude of the drug event reporting dynamic. The average drug and event reporting across reports equaled the event reporting rate multiplied by a fold change factor resulting in the effect size of dynamic ADE reporting.

**Table S1.**
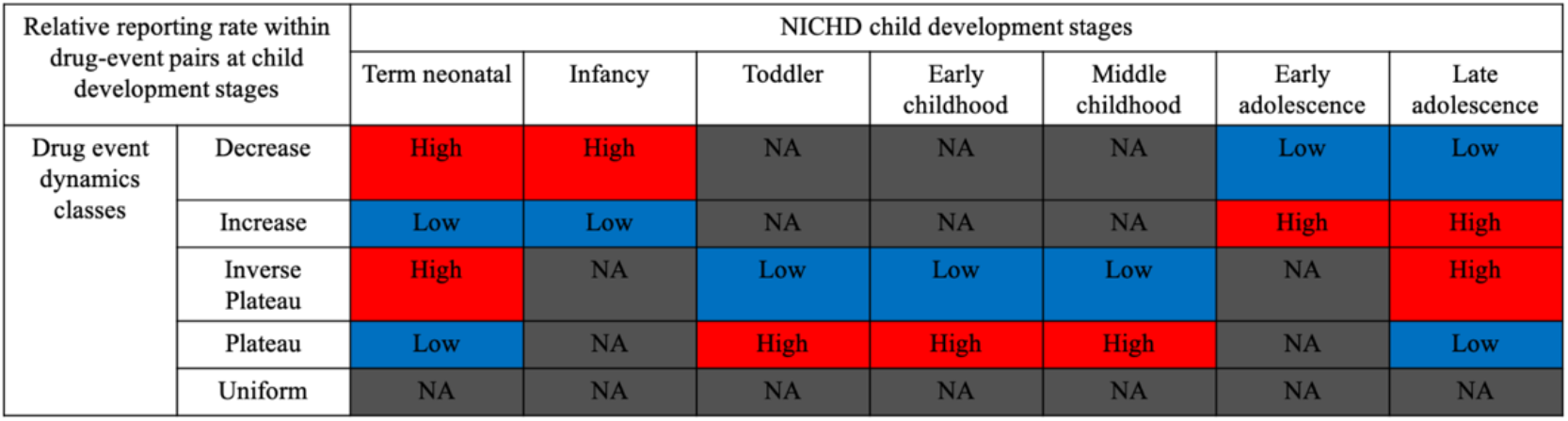
High and low drug, event reporting at child development stages for across each dynamics class.

**Table S2.** The number of drug-event pairs after the dynamics power analysis. Methods and scores had differences in drug reporting and effect size thresholds to identify drug, event pairs with atleast 80% power to differentiate scores between high and low reporting development stages.

| Number of drug-event pairs after the power analysis |  | Drug event dynamics class |  |  |  |
| --- | --- | --- | --- | --- | --- |
|  |  | Decrease | Increase | Inverse Plateau | Plateau |
| Score | PRR | 282 (56.4%) | 283 (56.6%) | 282 (56.4%) | 282 (56.4%) |
|  | PRR_90mse | 109 (21.8%) | 109 (21.8%) | 109 (21.8%) | 109 (21.8%) |
|  | GAM | 109 (21.8%) | 109 (21.8%) | 109 (21.8%) | 109 (21.8%) |
|  | GAM_90mse | 68 (13.6%) | 68 (13.6%) | 68 (13.6%) | 68 (13.6%) |

## Notes

### Competing Interest Statement

The authors have declared no competing interest.

### Funding Statement

The authors wish to acknowledge funding support from the National Institutes of General Medical Sciences (R35 GM1319050). N.P.G. and N.P.T. are both funded by R35 GM131905.

### Author Declarations

No IRB approval was needed. All data was publically available.

## References

1. Impicciatore P, Choonara I, Clarkson A, Provasi D, Pandolfini C, Bonati M. Incidence of adverse drug reactions in paediatric in/out-patients: a systematic review and meta-analysis of prospective studies. Br J Clin Pharmacol. 2001;52(1):77–83. doi:10.1046/j.0306-5251.2001.01407.x

2. Smyth RMD, Gargon E, Kirkham J, et al. Adverse drug reactions in children--a systematic review. PLoS One. 2012;7(3):e24061. doi:10.1371/journal.pone.0024061

3. Giangreco NP, Elias JE, Tatonetti NP. No population left behind: Improving paediatric drug safety using informatics and systems biology. Br J Clin Pharmacol. Published online December 17, 2020:bcp.14705. doi:10.1111/bcp.14705

4. Benjamin DK, Smith PB, Sun MJM, et al. Safety and Transparency of Pediatric Drug Trials. Arch Pediatr Adolesc Med. 2009;163(12):1080–1086. doi:10.1001/archpediatrics.2009.229

5. de Bie S, Ferrajolo C, Straus SMJM, et al. Pediatric Drug Safety Surveillance in FDA-AERS: A Description of Adverse Events from GRiP Project. Garattini S, ed. PLoS One. 2015;10(6):e0130399. doi:10.1371/journal.pone.0130399

6. Castro-Pastrana LI, Carleton BC. Improving pediatric drug safety: need for more efficient clinical translation of pharmacovigilance knowledge. J Popul Ther Clin Pharmacol = J la Ther des Popul la Pharmacol Clin. 2011;18(2):e76–88. http://journals.sagepub.com/doi/10.1038/jcbfm.2012.176

7. Crespi B. The evolutionary biology of child health. Proc R Soc B Biol Sci. 2011;278(1711):1441–1449. doi:10.1098/rspb.2010.2627

8. Yaffe S, Estabrook RW, Pitluck S, Davis JR. Rational Therapeutics for Infants and Children.; 2000. doi:10.17226/9816

9. Johnson T. The development of drug metabolising enzymes and their influence on the susceptibility to adverse drug reactions in children. Toxicology. 2003;192(1):37–48. doi:10.1016/S0300-483X(03)00249-X

10. Becker ML, Leeder JS. Identifying genomic and developmental causes of adverse drug reactions in children. Pharmacogenomics. 2010;11(11):1591–1602. doi:10.2217/pgs.10.146

11. de Graaf-Peters VB, Hadders-Algra M. Ontogeny of the human central nervous system: What is happening when? Early Hum Dev. 2006;82(4):257–266. doi:10.1016/j.earlhumdev.2005.10.013

12. Simon AK, Hollander GA, McMichael A. Evolution of the immune system in humans from infancy to old age. Proceedings Biol Sci. 2015;282(1821):20143085. doi:10.1098/rspb.2014.3085

13. Carvalho FS, Burgeiro A, Garcia R, Moreno AJ, Carvalho RA, Oliveira PJ. Doxorubicin-Induced Cardiotoxicity: From Bioenergetic Failure and Cell Death to Cardiomyopathy. Med Res Rev. 2014;34(1):106–135. doi:10.1002/med.21280

14. Guo H-L, Jing X, Sun J-Y, et al. Valproic Acid and the Liver Injury in Patients with Epilepsy: An Update. Curr Pharm Des. 2019;25(3):343–351. doi:10.2174/1381612825666190329145428

15. Moon YE. Paradoxical reaction to midazolam in children. Korean J Anesthesiol. 2013;65(1):2–3. doi:10.4097/kjae.2013.65.1.2

16. Kawakami Y, Fujii S, Ishikawa G, Sekiguchi A, Nakai A, Takase M. Valproate-induced polycystic ovary syndrome in a girl with epilepsy: A case study. J Nippon Med Sch. 2018;85(5):287–290. doi:10.1272/jnms.JNMS.2018_85-46

17. Cui N, Wu F, Lu W, et al. Doxorubicin-induced cardiotoxicity is maturation dependent due to the shift from topoisomerase IIα to IIβ in human stem cell derived cardiomyocytes. N. 2019;23(7):4627–4639. doi:10.1111/jcmm.14346

18. Park SO, Wamsley HL, Bae K, et al. Conditional Deletion of Jak2 Reveals an Essential Role in Hematopoiesis throughout Mouse Ontogeny: Implications for Jak2 Inhibition in Humans. PLoS One. 2013;8(3):1–14. doi:10.1371/journal.pone.0059675

19. Marret S, Mukendi R, Gadisseux JF, Gressens P, Evrard P. Effect of ibotenate on brain development: An excitotoxic mouse model of microgyria and posthypoxic-like lesions. J Neuropathol Exp Neurol. 1995;54(3):358–370. doi:10.1097/00005072-199505000-00009

20. Jiang X-L, Zhao P, Barrett J, Lesko L, Schmidt S. Application of Physiologically Based Pharmacokinetic Modeling to Predict Acetaminophen Metabolism and Pharmacokinetics in Children. CPT Pharmacometrics Syst Pharmacol. 2013;2(10):80. doi:10.1038/psp.2013.55

21. Krekels EHJ, Neely M, Panoilia E, et al. From pediatric covariate model to semiphysiological function for maturation: Part I-extrapolation of a covariate model from morphine to zidovudine. CPT Pharmacometrics Syst Pharmacol. 2012;1(1):1–8. doi:10.1038/psp.2012.11

22. Baldrick P. Juvenile animal testing in drug development -Is it useful? Regul Toxicol Pharmacol. 2010;57(2-3):291–299. doi:10.1016/j.yrtph.2010.03.009

23. Goulooze SC, Zwep LB, Vogt JE, et al. Beyond the Randomized Clinical Trial: Innovative Data Science to Close the Pediatric Evidence Gap. Clin Pharmacol Ther. 2020;107(4):786–795. doi:10.1002/cpt.1744

24. Brussee JM, Calvier EAM, Krekels EHJ, et al. Children in clinical trials: towards evidence-based pediatric pharmacotherapy using pharmacokinetic-pharmacodynamic modeling. Expert Rev Clin Pharmacol. 2016;9(9):1235–1244. doi:10.1080/17512433.2016.1198256

25. Christensen ML, Davis RL. Identifying the “Blip on the Radar Screen”: Leveraging Big Data in Defining Drug Safety and Efficacy in Pediatric Practice. J Clin Pharmacol. 2018;58(January):S86–S93. doi:10.1002/jcph.1141

26. Tatonetti NP. Translational medicine in the Age of Big Data. Brief Bioinform. 2019;20(2):457–462. doi:10.1093/bib/bbx116

27. Berlin JA, Glasser SC, Ellenberg SS. Adverse event detection in drug development: Recommendations and obligations beyond phase 3. Am J Public Health. 2008;98(8):1366–1371. doi:10.2105/AJPH.2007.124537

28. Etwel FA, Rieder MJ, Bend JR, Koren G. A Surveillance Method for the Early Identification of Idiosyncratic Adverse Drug Reactions. Drug Saf. 2008;31(2):169–180. doi:10.2165/00002018-200831020-00006

29. Star K, Sandberg L, Bergvall T, Choonara I, Caduff-Janosa P, Edwards IR. Paediatric safety signals identified in VigiBase: Methods and results from Uppsala Monitoring Centre. Pharmacoepidemiol Drug Saf. 2019;28(5):680–689. doi:10.1002/pds.4734

30. Evans SJW, Waller PC, Davis S. Use of proportional reporting ratios (PRRs) for signal generation from spontaneous adverse drug reaction reports. Pharmacoepidemiol Drug Saf. 2001;10(6):483–486. doi:10.1002/pds.677

31. Osokogu OU, Dodd C, Pacurariu A, Kaguelidou F, Weibel D, Sturkenboom MCJM. Drug Safety Monitoring in Children: Performance of Signal Detection Algorithms and Impact of Age Stratification. Drug Saf. 2016;39(9):873–881. doi:10.1007/s40264-016-0433-x

32. Hastie T, Tibshirani R, Friedman J. Elements of Statistical Learning. 2nd editio. Springer; 2008. http://web.stanford.edu/~hastie/pub.htm

33. Guisan A, Edwards TC, Hastie T. Generalized linear and generalized additive models in studies of species distributions: setting the scene. Ecol Modell. 2002;157(2-3):89–100. doi:10.1016/S0304-3800(02)00204-1

34. Yin Q, Wang J. The association between consecutive days’ heat wave and cardiovascular disease mortality in Beijing, China. BMC Public Health. 2017;17(1):1–10. doi:10.1186/s12889-017-4129-7

35. Tamayo-Uria I, Mateu J, Escobar F, Mughini-Gras L. Risk factors and spatial distribution of urban rat infestations. J Pest Sci (2004). 2014;87(1):107–115. doi:10.1007/s10340-013-0530-x

36. Fabiano V, Mameli C, Zuccotti GV. Adverse drug reactions in newborns, infants and toddlers: pediatric pharmacovigilance between present and future. Expert Opin Drug Saf. 2012;11(1):95–105. doi:10.1517/14740338.2011.584531

37. Saghir SA, Khan SA, McCoy AT. Ontogeny of mammalian metabolizing enzymes in humans and animals used in toxicological studies. Crit Rev Toxicol. 2012;42(5):323–357. doi:10.3109/10408444.2012.674100

38. Gunewardena SS, Yoo B, Peng L, et al. Deciphering the Developmental Dynamics of the Mouse Liver Transcriptome. Buratti E, ed. PLoS One. 2015;10(10):e0141220. doi:10.1371/journal.pone.0141220

39. Bohn J, Kortepeter C, Muñoz M, Simms K, Montenegro S, Dal Pan G. Patterns in spontaneous adverse event reporting among branded and generic antiepileptic drugs. Clin Pharmacol Ther. 2015;97(5):508–517. doi:10.1002/cpt.81

40. Bagattini F, Karlsson I, Rebane J, Papapetrou P. A classification framework for exploiting sparse multi-variate temporal features with application to adverse drug event detection in medical records. BMC Med Inform Decis Mak. 2019;19(1):1–21. doi:10.1186/s12911-018-0717-4

41. Zhao J, Papapetrou P, Asker L, Boström H. Learning from heterogeneous temporal data in electronic health records. J Biomed Inform. 2017;65:105–119. doi:10.1016/j.jbi.2016.11.006

42. Wood SN. Generalized Additive Models: An Introduction with R, Second Edition.; 2017. doi:10.1201/9781315370279

43. Helwig NE. Regression with ordered predictors via ordinal smoothing splines. Front Appl Math Stat. 2017;3(July):1–13. doi:10.3389/fams.2017.00015

44. Simpson D, Rue H, Riebler A, Martins TG, Sørbye SH. Penalising model component complexity: A principled, practical approach to constructing priors. Stat Sci. 2017;32(1):1–28. doi:10.1214/16-STS576

45. Williams K, Thomson D, Seto I, et al. Standard 6: Age groups for pediatric trials. Pediatrics. 2012;129(SUPPL. 3):S153–S160. doi:10.1542/peds.2012-0055I

46. Kass-Hout TA, Xu Z, Mohebbi M, et al. OpenFDA: An innovative platform providing access to a wealth of FDA’s publicly available data. J Am Med Informatics Assoc. 2016;23(3):596–600. doi:10.1093/jamia/ocv153

47. Giangreco NP. ngiangre/openFDA_drug_event_parsing: Repository for python notebooks and scripts for parsing the json-formatted drug event reports collected by the FDA. Published online 2021. doi:https://zenodo.org/badge/latestdoi/246321623

48. OHDSI. Athena. Accessed January 17, 2021. https://athena.ohdsi.org/search-terms/start

49. Buja BYA, Hastie T, Tibshirani R. Linear Smoothers and Additive Models. 2016;17(2):453–510. https://scholar.google.com/scholar_lookup?journal=The+Annals+of+Statistics&title=Linear+smoothers+and+additive+models&author=A+Buja&author=T+Hastie&author=R+Tibshirani&publication_year=1989&pages=453-510&

50. Hastie T, Tibshirani R. Generalized additive models for medical research. Stat Methods Med Res. 1995;4(3):187–196. doi:10.1177/096228029500400302

51. Osokogu OU, Fregonese F, Ferrajolo C, et al. Pediatric drug safety signal detection: a new drug-event reference set for performance testing of data-mining methods and systems. Drug Saf. 2015;38(2):207–217. doi:10.1007/s40264-015-0265-0

52. Giangreco N. GRiP_pediatric_ADE-reference_set. Published 2020. Accessed October 21, 2020. https://github.com/ngiangre/GRiP_pediatric_ADE-reference_set

53. Giangreco NP. ngiangre/evaluating_ontogenic_ade_risk. Published online 2021. doi:10.5281/zenodo.4585585

